# COVID-19: Impact on the health and wellbeing of ex-serving personnel (Veterans-CHECK) protocol paper

**DOI:** 10.1101/2020.09.02.20186577

**Authors:** Marie-Louise Sharp, Danai Serfioti, Margaret Jones, Howard Burdett, David Pernet, Lisa Hull, Dominic Murphy, Sharon Stevelink, Simon Wessely, Nicola T Fear

**Author notes:** Joint First Authors (Marie-Louise Sharp, Danai Serfioti).

## Abstract

**Introduction:** We will use a sub-sample of a current longitudinal study to investigate the impact of COVID-19 on the health and wellbeing of ex-service personnel in the UK. The study will provide evidence for the UK Office of Veterans’ Affairs (OVA), UK stakeholders supporting the ex-service community, and evidence to inform our international counterparts working with ex-service communities in allied countries regarding the impact of COVID-19 on the health and wellbeing of ex-service personnel.

**Methods and analysis:** Participants were eligible to participate if they lived in the UK, had Regular service history from the UK Armed Forces and had previously completed the King’s Centre for Military Health Research (KCMHR) Health and Wellbeing survey between 2014–2016. Participants who met these criteria were recruited through email to take part in an online questionnaire. The study provides additional quantitative longitudinal data on this sub-sample. Data are being collected June 2020-September 2020. Specific measures are used to capture participants’ COVID-19 experiences, health and wellbeing status and lifestyle behaviours. Other key topics will include questions regarding the impact of COVID-19 pandemic on employment, finances, volunteering, charitable giving, accommodation and living arrangements, help-seeking behaviours, as well as any potential positive changes during this period.

**Ethics and Dissemination:** Ethical approval has been gained from King’s College London Research Ethics Committee (Ref: HR-19/20–18626). Participants were provided with information and agreed to a series of consent statements before enrolment. Data are kept on secure servers with access to personally identifiable information limited. Findings will be disseminated to the OVA, UK ex-service stakeholders and international research institutions through stakeholder meetings, project reports and scientific publications.

**Strengths and limitations of this study:**

- Strengths include recruitment from a population where underlying characteristics are known, and longitudinal data is held on their health and wellbeing.
- There has been rapid roll-out of the survey to ensure relevance for participants’ COVID-19 experiences and use of validated measures for mental health and wellbeing outcomes.
- Study limitations include recruitment from a specific cohort; hence the study cannot comment on the impact of COVID-19 in other veteran populations.

## Introduction

### Context

The COVID-19 pandemic raises many questions not only on biological, but also behavioural, emotional and social responses to a global threat with unknown long-term health impacts (PHOSP-COVID, 2020). Key questions include how to help people maintain unusual behaviours like social distancing or self-isolation or quarantine over many weeks, how best to provide key messages to promote health, and how to minimise the psychological and physical harms of the pandemic – including the impact of isolation, traumatic experience of illness and bereavement, and longer term economic threats (GOV.UK, 2020, LGA, 2020). There are many policy relevant questions for national and international health responses to COVID-19 concerning the general population as well as specific groups of individuals that may have different risk profiles or behaviours, such as black, ethnic and minority groups (Aldridge et al., 2020), older age or vulnerable adults (Brooke and Jackson, 2020), and in the specific case of our study, the ex-service population.

As identified by Davis et al. (2020), to be able to engage with these questions, studies are needed that can rapidly collect data, engage with participants longitudinally, utilising web-based platforms for recruitment and data collection that avoids face-to-face contact in pandemics, whilst needing research that samples from populations where characteristics are known to minimise biases found in convenience samples.

The ex-service population are an estimated population of 2.4 million in Great Britain making up 5% of household residents aged 16 and over (MOD, 2019). Ex-service personnel are a valuable population of interest to Government and the nation, because of commitments enshrined in the Armed Forces Covenant to ensure the Armed Forces community is not disadvantaged compared to the general population, due to their Service (MOD, 2011). Through the KCMHR Health and Wellbeing Study of UK Armed Forces, a large proportion of participants have been followed up since 2003, with three waves of data collected on their health and wellbeing over the period 2003–2016 (Hotopf et al., 2006, Fear et al., 2010, Stevelink et al., 2018). This study focuses on ex-service personnel in this survey, who at a minimum participated in the third wave of data collection of the study 2014–2016. This continuing data will provide evidence in trends of health outcomes of this population.

### Current knowledge

This pandemic is unprecedented and is without previous historical data. A recent paper published by leading academics, funders and people with lived experiences (Holmes et al., 2020), identifies that research must investigate as a priority the adverse effects of the pandemic on the mental health of different populations to inform public health and policy responses.

Recently published COVID-19 studies in the UK general population suggest the pandemic is having negative impacts in terms of inequality (Wright et al., 2020), self-harm and suicidal ideation (Iob et al., 2020) and potential distress caused by quarantine (Brooks et al., 2020). We currently have little health and wellbeing information on important sub-groups within the UK general population and their experience of the COVID-19 pandemic.

Ex-serving military personnel provide an important population to study, as their attitudes and behaviours may differ from the civilian population during the COVID-19 pandemic. In contrast to the general population (excluding emergency responders), military personnel are trained to demonstrate readiness and resilience in the face of the increasingly fast-paced nature of warfare operations and stressful environments, being able to maintain optimal cognitive and physical performance necessary for success (Nindl et al., 2018). Having left the military, ex-service personnel remain a unique, multifaceted population with a distinct culture that includes values, selfless duty, codes of conduct, and obedience to command (Olenick et al., 2015). However, transition experiences from military to civilian life vary greatly among ex-service personnel and part of this population face major barriers, including mental and physical health issues along with help-seeking barriers due to their needs (Stevelink et al., 2018, Murphy and Busuttil, 2019). Additional key barriers facing the ex-service community can include employment (Carolan, 2019), finances and housing (RBL, 2014), alcohol misuse (Irizar et al., 2020) and gambling (Roberts et al., 2020).

The Veterans-CHECK study will address some of the unanswered questions about the impact of the COVID-19 pandemic on the ex-service population. The study will provide evidence for the Government, OVA and ex-service stakeholders, such as Armed Forces charities, to help shape policy and support services to minimise any negative impact on the ex-service community, and as such fulfil commitments enshrined within the Armed Forces Covenant. This study will be able to compare outcomes with previous study data collected in 2014–2016 and will be able to compare outcomes with COVID-19 studies in the general population, as well as with international ex-service populations in allied countries. The intent is that these individuals would also be followed up in more waves of data collection, and therefore it is possible these outcomes will be able to be compared into the future to assess impact and trends.

### Research aims

This study is an exploratory study and therefore does not posit hypotheses. The primary aim of this study is to investigate the current experiences and impact of the COVID-19 pandemic on the health, wellbeing and lifestyle behaviours of ex-serving military personnel. This is a quantitative project which will focus on these key areas of interest, aiming to identify potential emerging issues in this population and psychological indicators of resilience and vulnerability.

Secondary aims include comparisons with health and lifestyle behaviours of ex-serving personnel before the COVID-19 pandemic, and with groups in the general population, for example other occupational groups, such as KCL or NHS staff (i.e., using the pseudonymised datasets of KCL CHECK (https://kcl-check.org/) and NHS CHECK (https://www.nhscheck.org/) research projects).

## Method and Analysis

### Study protocol

#### Sample population and eligibility

Participants will be recruited from the KCMHR Health and Wellbeing survey. This is a large-scale ongoing investigation of the physical and psychological health and wellbeing of UK Armed Forces personnel from all three Services (regulars and reservists) and includes personnel, some of whom were first surveyed before the conflicts in Iraq (Rona et al., 2006), and the majority during and after the conflicts in Iraq and Afghanistan. There have been three phases of data collection since the start of the Iraq conflict, Phase 1: 2004 – 2006, Phase 2: 2007 – 2009 and Phase 3: 2014 – 2016 (Hotopf et al., 2006, Fear et al., 2010, Stevelink et al., 2018). Approximately 18,000 have taken part in the survey since it began. For this study, only participants who have completed Phase 3, left the UK Armed Forces, had Regular service, reside within the UK, have consented to further follow up and have provided a valid email address will be sent an invitation to participate. From our cohort study, recruitment emails will be sent out to N = 4017 of individuals we believe meet this eligibility criteria.

Mandatory questions at the beginning of the survey determine eligibility. Participants will be excluded if they do not live in the UK, are still serving in the UK Armed Forces or have only ever served as a reservist. Participants will also be excluded if they are unable to access the internet to complete the survey, unable to access an email address to facilitate survey registration and receive survey links, or if we are unable to contact them – identified by bounce back emails from redundant email addresses.

The research will be conducted online. One invite email will be sent to participants in June 2020 and up to three email reminders will be sent in June, July and August 2020. The email invite includes information on the study, a participant information sheet (PIS), links to the study website, and a unique, personal link for each participant to take part in the study through the online questionnaire. Participants will be asked to complete a questionnaire in their personal settings (e.g., home) through REDCap (REDCap). REDCap is a secure web application for building and managing online surveys and databases. Consent will be completed online on the REDCap platform. Please note that we will not provide participants with an option to complete a paper questionnaire due to the COVID-19 infection risk. The online questionnaire will take approximately 15–20 minutes to complete.

The PIS provides detailed information on the study and answers to frequently asked questions. The PIS is attached to the participants own email invite and can be accessed from the consent page of the questionnaire, and the study website. The options regarding refusal to take part or withdrawal from the study are explained in the PIS. If potential participants do not wish to take part or they change their mind after completing the survey, they are informed on how to do so. Participants can withdraw by stopping completion of the online survey. Participants can email the study team using the dedicated study email address to indicate they are no longer willing to participate in the study. Participants can withdraw their questionnaire responses after they have taken part, up until the end of data collection. Data collection will close September 2020.

### Measures

Specific measures will be used to capture participants’ health and wellbeing status and lifestyle behaviours:

- 12-item General Health Questionnaire (GHQ-12) to screen for general (non-psychotic) psychiatric morbidity (Goldberg and Blackwell, 1970).
- 20-item PCL-5 to explore post-traumatic stress disorder (Weathers et al., 2013).
- 7-item Anger DAR-R to assess problematic anger (Forbes et al., 2004).
- 14-item Warwick-Edinburgh Mental Well-being Scale (WEMWBS) to enable the monitoring of subjective well-being and psychological functioning compared to the general population (all items are worded positively and address aspects of positive mental health) (Tennant et al., 2007).
- 10-item Alcohol Use Disorder Identification Test (AUDIT) to measure alcohol consumption and misuse (Babor et al., 2001).
- 3-item Clinical Interview Schedule-Revised (CIS-R) suicidal ideation/self-harm (Lewis et al., 1992)
- 3-item UCLA Loneliness Scale to measure feelings of isolation and loneliness (Hughes et al., 2004).
- 6-item Perceived Social Support Questionnaire (F-SozU) to measure participants’ perceptions of social support during the COVID-19 pandemic (Kliem et al., 2015)
- 3-item Problem Gambling Severity Index, short version (PGSI Mini), to investigate risk behaviour in problem gambling (Williams and Volberg, 2012)

Other key topics will include questions regarding the impact of COVID-19 pandemic on employment, finances, volunteering, charitable giving, lifestyle behaviours, accommodation and living arrangements, help-seeking behaviours, as well as any potential positive changes during this period

### Statistical methods

Response rates will be calculated based on our administrative records from the previous KCMHR 2014–2016 survey on numbers of eligible participants. Information from the survey will be summarised with appropriate summary statistics (e.g. mean and SD for continuous measures; frequencies and proportions for categorical measures). All scales will be scored according to published instructions. Missing items on the GHQ, PCL-5 and AUDIT will be handled according to the protocol used in the previous KCMHR surveys, i.e. the lowest value is imputed if fewer than four items are missing on the measure. Differences in the composition of survey respondents will be compared to the overall eligible participant group and will be summarised. The assessment of the characterisation of survey respondents will draw on administrative records from the KCMHR survey 2014–2016 for ex-service personnel such as age, gender, rank and service branch.

The prevalence of probable mental disorders and wellbeing outcomes will be summarised using descriptive statistics. Associations between COVID-19 experiences and mental health will be tested using linear and logistic models as appropriate, with adjustments for measured confounders.

### Data storage and security

Individuals who participate will be automatically assigned an ID number once their informed consent is obtained, allowing their online survey data to be held pseudonymously.

Online survey responses will be collected using REDCap survey software which will be hosted by King’s College London, on servers located in the UK. Data required for analysis will be downloaded in pseudonymous form and stored on KCL OneDrive. Any data held electronically will only be accessible by specified members of the research team who have been given authorisation to access the database. Access to the server is via assignment as an authorised user (a research team staff member who has a King’s ID number and a password which complies with the King’s password policy(i.e. containing a combination of numbers, letter and symbols). Authorised users are reviewed regularly and updated as needed (e.g. when a member of the research team leaves). Data will be stored separately from personal information. No identifying information will be presented in any reports or publications.

### Data management and oversight

All research staff are provided with training regarding the General Data Protection Regulation (GDPR), KCL standards for handling data and Medical Research Council (MRC) data protection training.

Data required for analysis will be downloaded in pseudonymised form and stored on KCL OneDrive. Any interactions with REDCap will be logged and audited by a designated researcher. We will use a valid Data Protection and Security Toolkit from KCMHR at King’s College London and will follow this Toolkit during rollout. Data will be kept by KCMHR for twenty years as we may use the pseudonymous data in future research.

### Patient and public involvement

Veterans who sit on our research board tested the design and flow of the questionnaire and offered advice on outcome measures. As a result of this feedback, a new section on the questionnaire was added to measure volunteering and charitable giving. This was added to reflect potential positive outcomes associated with the COVID-19 pandemic on veterans– behaviours. This addition was based on feedback from veterans that during times of crisis, veterans often wanted to contribute and give back to their communities as a learnt part of their service culture. Results of the study will be disseminated to study participants through the KCMHR newsletter, through our social media outlets and through our stakeholders that represent veteran communities.

### Ethical and safety considerations

Ethical approval has been gained from King’s College London Research Ethics Committee (Ref: HR-19/20–18626).

### Confidentiality

To ensure participant confidentiality. Data will be stored separately from personal information (individuals who participate will be automatically assigned an ID number once their informed consent is obtained, allowing their online survey data to be held pseudonymously). No identifying information will be presented in any reports or publications. Aggregated data will be made available in reports for the OVA and future publications. Cells based on small numbers (i.e., n < 5) will be supressed. Pseudonymised datasets may be shared with other research institutions, but personal identifiable data will never be shared with other researchers outside of KCL.

### Participant safety

To ensure participant safety when taking part in this study, the nature of the questionnaire is clearly explained on the PIS. The study webpage includes information about the study, the PIS, and resources signposting people to support services and helplines are available to revisit at any time. Participants are able to stop participating in the survey at any point and can skip questions (apart from the consent and eligibility questions at the beginning of the survey). Support information is made available during the survey for participants who recognise that they are feeling distressed (e.g., after the suicidal ideation/self-harm section). After participants have completed the survey, participants are offered a variety of resources signposting them to support services and helplines (including Armed Forces charities, NHS, charities or further guidance for urgent help for ex-serving personnel).

It is highlighted in the PIS that participants can contact the research team at a dedicated study email and mobile telephone number if they have any questions regarding the conduct of study (i.e., information that is covered in the Information Sheet). An automatic reply is set up, informing participants who email us that we will be able to respond to their emails within the next two working days, highlighting that the inbox will not be monitored outside regular office hours (i.e., 09:00 to 17:00, Monday to Friday). The automatic reply will include links to the study webpage and the PIS, which include information about the study and resources signposting participants to support services and helplines. In the unlikely event someone contacts us to ask for advice regarding their mental health, they will be directed to seek medical advice via their GP or in an emergency A&E.

### Participant burden

The questionnaire has been designed to limit the burden of questions both in volume and experience negotiating the questionnaire. For example, the study has focused on measures that are essential or comparable to other COVID-19 studies to maximise comparability to other populations. The questionnaire survey was tested by the KCMHR research team and veterans to ensure efficient flow of questions and appropriate language. To reduce the participant burden, screening questions were applied were possible (e.g. AUDIT, PGSI). Only participants who respond positively to the screening question(s) are invited to complete the full measures.

## Dissemination

Results will be presented at stakeholder meetings and at the Veterans Mental Health Conference 2021 at King’s College London. Results will be written up in academic papers and submitted for peer reviewed publication. A study report will be presented to the OVA, and academic publications will be disseminated through KCL communications channels in social media, press briefings and academic conferences.

## Funding and roles

This project has been funded by the OVA, Cabinet Office, UK Government. All investigators are employed by King’s College London. Decisions regarding study design, and the collection, and management of data were made by the named investigators. Decisions regarding the analysis, interpretation and writing of reports will be carried out by the named investigators. The investigators have regular meetings to agree study design and analysis decisions. The decision to submit reports to publication will remain under the ultimate authority of the principal investigators. The views expressed in this and future papers will be those of the authors and not necessarily those of the King’s College London or the OVA.

## Conclusions

Veterans-CHECK is an online study aimed at investigating the impact of COVID-19 on the ex-service population of the UK. The research entails data collection June-September 2020 focused on COVID-19 experiences, but forms part of a longitudinal study, recruiting its sample population from the KCMHR Health and Wellbeing survey that has followed up participants since 2003. One aim of this study is to give the OVA and Armed Forces stakeholders evidence on which to make assessment of need in the veteran community and provide policy and practical support where necessary. The study strengths include recruitment from a population where underlying characteristics are known and longitudinal data held on health and wellbeing, rapid roll-out, and use of validated measures for mental health and well-being outcomes. Study limitations include recruitment from a specific cohort; hence the study cannot comment on the impact of COVID-19 on other veteran populations. The study is planned to last 9 months.

## Data Availability

Data is currently being collected. Data will be processed in accordance with the General Data Protection Regulation (GDPR) and the Data Protection Act 2018. We will not make any record-level data publicly accessible because we need to protect the confidentiality and security of the individual cohort members. You are welcome to contact us with proposals for collaborative research, which the investigators will consider on a case-by-case basis, and which will only occur as part of a legal collaborative agreement and after the collaborator has put in place the relevant research ethics, data protection, and data access approvals.

## Acknowledgments

This paper represents independent research funded by the Office of Veterans’ Affairs (OVA), UK. The views expressed are those of the authors and not necessarily those of King’s College London or the OVA. Thank you to KCMHR colleagues and veterans who helped to edit and shape the survey questionnaire.

## Authors’ Contributions

MLS, DS, MJ, HB, DP, LH, DM, SM, NF were involved in the original concept and design of the study. NF and SM have overseen the conduct of all aspects of the study. SS provided resources to assess choice of outcome measures and provided the framework utilised for this protocol manuscript. MLS led on the formulation of questionnaire outcome measures, with substantial contributions from DS, MJ, HB, DP, LH, DM, SM and NF in shaping the final questionnaire. DP led on online survey design, format and flow. DS led on the ethics submission with substantial contributions from MLS, MJ, HB, DP, LH, DM, SM and NF. MJ, HB and LH led on the design of participant materials including the participant invite and information sheet with input from all authors. MJ and HB led on the data analysis plan. MLS and DS led the writing of the protocol paper, with drafting and revision input from all named authors. MLS, DS, MJ, HB, DP, LH, DP, DM, SS, SM, NF have all approved the final version of this paper and accept accountability for all aspects of the work.

